# *FADS2* Indel polymorphism rs66698963 predicts colorectal polyp prevention by the *n*-3 fatty acid EPA

**DOI:** 10.1101/2023.10.28.23297412

**Authors:** Ge Sun, Yan Ning Li, John R Davies, Robert Block, Kumar S D Kothapalli, J Thomas Brenna, Mark A Hull

**Author notes:** **Corresponding author:** M A Hull, Division of Gastrointestinal and Surgical Sciences, Leeds Institute of Medical Research, St James’s University Hospital, University of Leeds, Leeds, LS9 7TF, United Kingdom. **Bold** denotes Authors that contributed equally to the study.

## Abstract

**Importance:** A precision medicine approach to identify who would benefit from supplementation with the *n*-3 highly unsaturated fatty acid (HUFA) eicosapentaenoic acid (EPA) for colorectal cancer prevention has not been reported. A *fatty acid desaturase 2 (FADS2)* insertion-deletion (Indel) polymorphism (rs66698963) controls levels of the *n*-6 HUFA arachidonic acid (AA), which drives intestinal tumorigenesis and which is antagonized by EPA.

**Objective:** We tested the hypothesis that the *FADS2* Insertion (I) allele, which is associated with elevated AA levels, predicts those individuals who display colorectal polyp risk reduction by EPA.

**Design:** Secondary analysis of the randomized, placebo-controlled, 2×2 factorial seAFOod polyp prevention trial of EPA 2g daily and aspirin 300mg daily, stratified for *FADS2* Indel genotype.

**Setting:** Colonoscopy surveillance 12 months after clearance screening colonoscopy, in the English Bowel Cancer Screening Programme (BCSP).

**Participants:** A predominantly White European, male cohort (mirroring the BCSP colonoscopy demographic). 528 trial participants with colonoscopy data and a *FADS2* Indel genotype from the original randomized trial population of n=707.

**Main Outcome(s) and Measure(s):** Total (adenomatous and serrated) colorectal polyp risk associated with EPA or aspirin compared with its respective placebo. Presence of at least one I allele and an interaction term (at least one I allele x active intervention) were co-variates in negative binomial regression models.

**Results:** EPA use, irrespective of *FADS2* Indel genotype, was not associated with reduced total colorectal polyp number (incidence rate ratio [IRR] 0.92, 95% confidence interval 0.74,1.16), mirroring the original seAFOod trial analysis. However, the presence of at least one I allele identified EPA users with a significant reduction in colorectal polyp number (IRR 0.50 [0.28, 0.90]), unlike aspirin for which there was no evidence of an interaction. Similar findings were obtained for analysis of the polyp detection rate (% of individuals with at least one polyp).

**Conclusions and Relevance:** The *FADS2* Indel I allele identifies individuals who display colorectal polyp prevention efficacy of EPA, with a similar effect size to aspirin. Assessment of rs66698963 as a therapeutic response biomarker in other populations and healthcare settings is warranted.

**Trial Registration:** The seAFOod polyp prevention trial and STOP-ADENOMA project - ISRCTN05926847.

**Key points:** *Question:* Does a functional *fatty acid desaturase 2 (FADS2)* insertion-deletion (Indel) polymorphism (rs66698963) predict colorectal polyp prevention efficacy of eicosapentaenoic acid (EPA)?

*Findings:* In 528 participants of the 2 × 2 factorial seAFOod polyp prevention trial of the *n*-3 highly unsaturated fatty acid (HUFA) EPA and aspirin, who had both colonoscopy outcome and Indel genotype data, a gene (I allele carrier) x treatment interaction identified individuals for whom EPA significantly reduced colorectal polyp number by approximately 50% (a similar effect size to aspirin).

*Meaning:* Further evaluation of a precision medicine approach using the *FADS2* Indel polymorphism rs66698963 as a therapeutic response biomarker for cancer and cardiovascular disease prevention by *n*-3 HUFAs is warranted.

## Introduction

The *n*-3 highly unsaturated fatty acid (HUFA) C20:5*n*-3 eicosapentaenoic acid (EPA) has anti-colorectal cancer (CRC) activity [1]. EPA 2g daily for 6 months reduced rectal adenomatous polyp number and size by approximately 20% compared with placebo in familial adenomatous polyposis patients [2]. However, the efficacy of the same dose of EPA was modest, and did not reach statistical significance (incidence rate ratio [IRR] 0.91, 95% confidence interval [CI] 0.79, 1.05), for reduction in colorectal polyp number in the seAFOod 2×2 factorial polyp prevention trial of EPA and aspirin 300mg daily in patients undergoing colonoscopy surveillance after removal of multiple ‘sporadic’ colorectal polyps [3].

The mechanism of the anti-neoplastic activity of EPA is uncertain, but the strongest evidence suggests that EPA antagonizes pro-tumorigenic signalling by competing with its *n*-6 HUFA homologue C20:4*n*-6 arachidonic acid (AA) as a substrate for cyclooxygenase (COX)-dependent synthesis of pro-tumorigenic prostaglandin (PG) E_2_ [4]. The rate-limiting step for endogenous AA synthesis from dietary C18:2*n*-6 linoleic acid (LA) is Δ5-desaturation by fatty acid desaturase 1 (FADS1) [5-6]. We have previously characterized a functional 22 base pair (bp) insertion-deletion (Indel) polymorphism (rs66698963) in intron 1 of the neighbouring *FADS2* gene, which is 137bp downstream of a sterol-response element [7]. The Indel is evolutionarily selective, probably based on ancestral diet, and controls circulating AA levels via FADS1 in free-living humans across several diverse populations [7-9]. Homozygosity for the insertion (I) allele (a 22bp repeat) has the effect of increasing circulating AA levels by up to 50% compared with deletion (D) allele (a single 22bp copy) homozygotes [8-9].

We hypothesized that *FADS2* Indel polymorphism rs66698963 predicts colorectal polyp prevention efficacy of EPA based on competition between EPA and elevated bioavailability of AA in I allele carriers.

## Methods

The randomized, double-blind, placebo-controlled, 2×2 factorial seAFOod polyp prevention trial, as well as secondary genetic and lipid mediator analyses, have been reported [3, 10-13]. Details of rs66698963 genotyping, as well as red blood cell (RBC) membrane HUFA and plasma oxylipin measurements, are in Supplementary Methods.

We investigated the effect of EPA and aspirin treatment on total (adenomatous and serrated) colorectal polyp number at trial exit colonoscopy using Poisson and negative binomial regression models (see Supplementary Methods). Data are reported as the incidence rate ratio (IRR) and 95% confidence interval (CI). Logistic regression examined factors predicting the polyp detection rate (PDR; % individuals with one or more colorectal polyps). Consistent with previous primary and secondary seAFOod polyp prevention trial analysis, we included repeat baseline colonoscopy and adjusted for the research colonoscopy site in all models [3, 10-11]. Male sex was a covariate given the consistent relationship with increased colorectal polyp risk in previous studies [14]. All analyses were performed ‘at the margins’ of the 2×2 factorial trial, i.e., each active intervention (EPA or aspirin) was compared to its respective placebo, regardless of the other intervention [3]. Presence of at least one I allele was included as a co-variate, as well as an interaction term (at least one I allele x active intervention). Finally, a combined model was derived that included EPA and aspirin use as separate terms given that *post-hoc* trial analysis suggested that combination treatment was associated with reduced colorectal polyp recurrence compared with single agent use [13].

## Results

A FADS2 Indel genotype was available for 630 seAFOod trial participants (Supplementary Figure 1). There were 64 (10.2%) I/I homozygotes in this predominantly White European cohort. Given the small number of I/I homozygotes, we combined I homozygotes and heterozygotes (I/D) for comparison with D/D individuals (Supplementary Methods).

Consistent with previous reports [7, 9], individuals with at least one I allele exhibited a significantly higher RBC membrane AA/LA ratio compared to participants with a D/D genotype (median 1.10 [interquartile range 0.75-1.39] *versus* 1.01 [0.68-1.28]; p=0.008, Figure 1A). Trial participants with at least one I allele also had a higher plasma concentration of the aspirin-induced AA metabolite 15-hydroxyeicosatetraenoic acid (mean 0.79±1.43 [standard deviation]) than D/D individuals (0.28±1.11; p=0.03) after treatment for 6 months, compatible with increased substrate bioavailability of AA in I allele carriers (Figure 1B).

**Figure 1:**
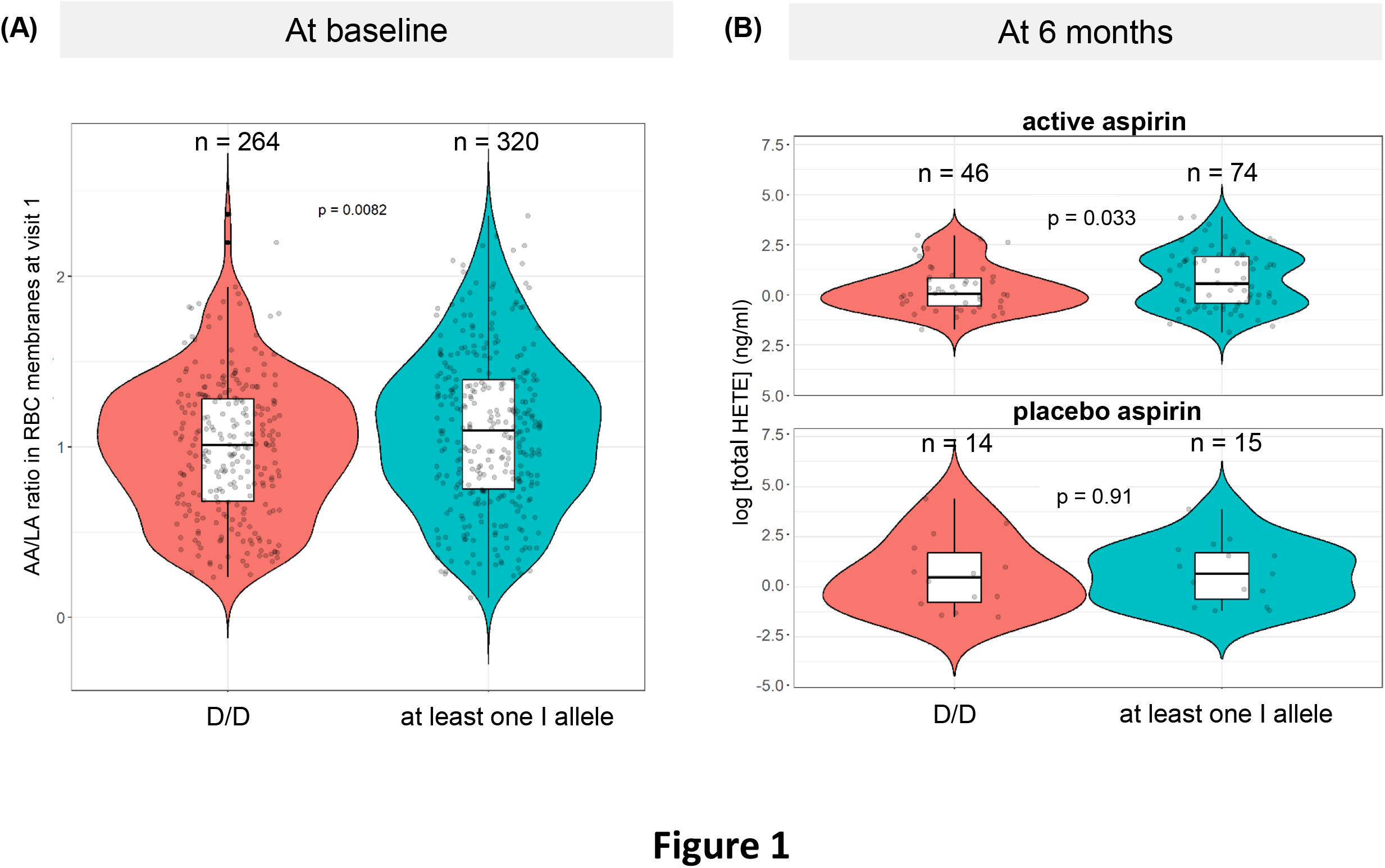
Comparison of the AA/LA ratio in red blood cell membranes (A) and plasma total 15-HETE concentration (B) according to FADS2 Indel genotype. (A) Violin plot, with embedded box and whisker plot and individual data points, of baseline (seAFOod trial visit 1) AA/LA ratio values in 584 trial participants with an Indel genotype and available red blood cell (RBC) HUFA data [3]. The AA/LA ratio was calculated using % levels of AA and LA of total fatty acids in red blood cell membranes measured by liquid chromatography-mass spectrometry [3]. P=0.008 for the difference between I carriers and D/D homozygotes (Wilcoxon test). (B) Violin plot, with embedded box and whisker plot and individual data points, of the plasma total (sum of *R*- and *S*-enantiomers) 15-HETE concentration at seAFOod trial visit 4 after treatment with active aspirin or placebo aspirin for 6 months [12]. P=0.033 for the difference in plasma total 15-HETE concentration between I carriers and D/D homozygotes in the aspirin group only (t-test).

A FADS2 Indel genotype and full colonoscopy outcome data were available for 528 seAFOod trial participants (Supplementary Figure 1). Clinical characteristics were similar to the original n=707 seAFOod trial population, for whom trial data were available (Supplementary Table 1). The distribution of FADS2 Indel genotypes was well balanced across the respective active *versus* placebo intervention groups (% I carrier for active EPA [52.8%] *vs* placebo EPA [57.7%]; active aspirin [56.6%] *vs* placebo aspirin [54.0%]).

EPA use, irrespective of FADS2 Indel genotype, was not associated with reduced total colorectal polyp number in the 528 individuals with a FADS2 Indel genotype (IRR 0.92 95%CI 0.74, 1.16) inside a Poisson regression model, mirroring the primary analysis of the seAFOod trial [3]. In the negative binomial regression model that included Indel genotype, EPA use was also not associated with any significant change in colorectal polyp risk (IRR 1.29 [0.84, 2.00]; Figure 2A). However, the presence of at least one I allele identified EPA users with a significant reduction in colorectal polyp number (IRR 0.50 [0.28, 0.90]; Figure 2A). Consistent with higher AA levels and elevated tumorigenic risk, at least one I allele was associated with a trend towards increased colorectal polyp risk (IRR 1.40 [0.94, 2.11]; Figure 2A).

**Figure 2:**
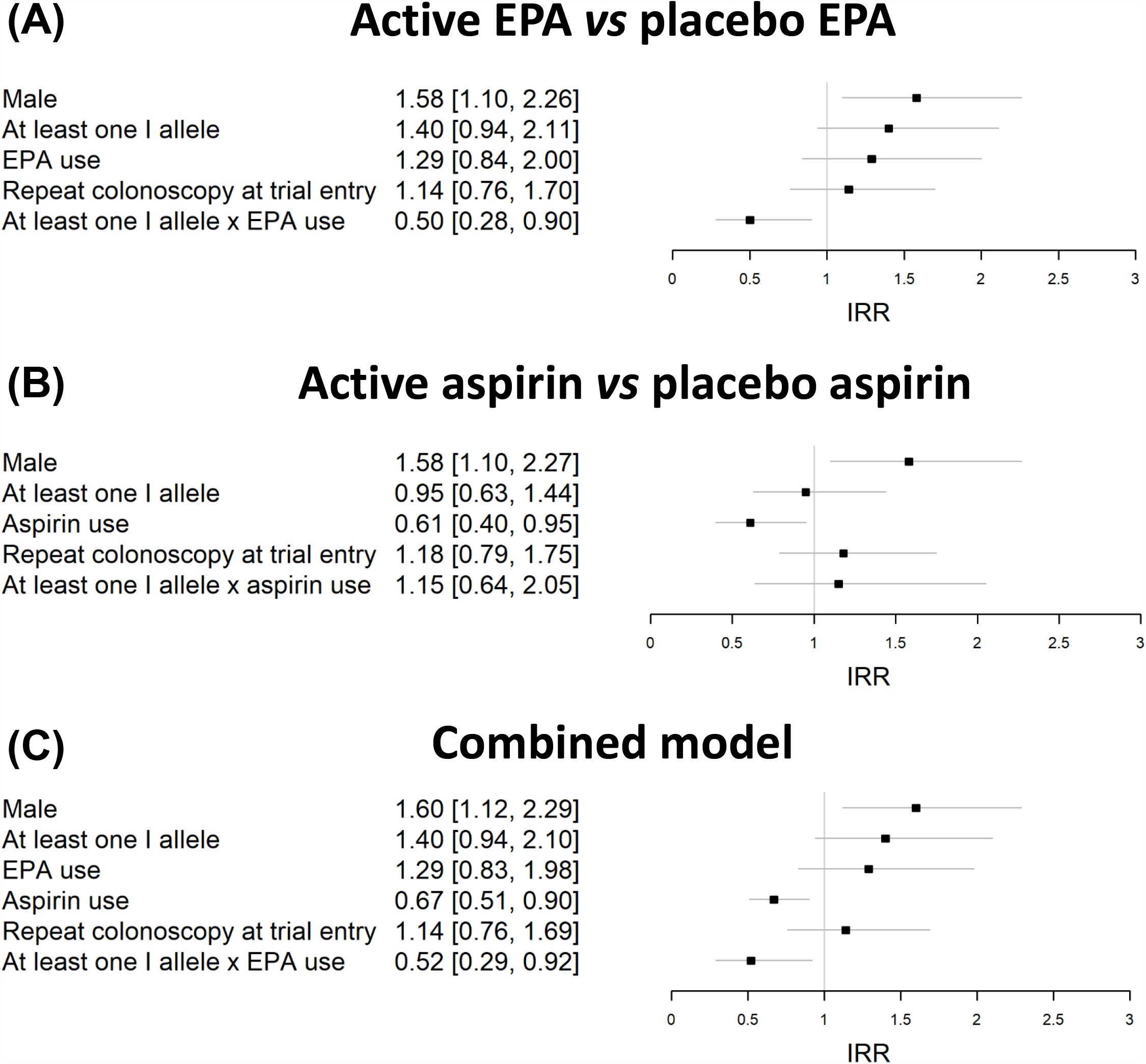
Multivariate models investigating the interaction between EPA or aspirin treatment, and FADS2 Indel genotype (I carrier status) for colorectal polyp number. Forest plots with the incidence rate ratio (IRR) and 95% confidence interval (horizontal lines) for co-variates for ‘at the margins’ comparison of (A) active EPA *vs* placebo EPA, (B) active aspirin *vs* placebo aspirin, as well as (C) a combined model including both treatments as independent co-variates.

By contrast, the regression model according to aspirin use demonstrated the polyp prevention efficacy of aspirin (IRR 0.61 [0.40, 0.95]; Figure 2B), but there was no significant interaction between Indel genotype and aspirin use, as expected if the primary mechanism of action of aspirin is cyclooxygenase inhibition, as opposed to limiting substrate (AA) availability (Figure 2B). The combined model with EPA and aspirin use as separate co-variates did not materially change the key finding that EPA use in trial participants with at least one I allele displayed lower colorectal polyp risk (IRR 0.52 [0.29, 0.92]), comparable to the effect size associated with aspirin use (Figure 2C).

Logistic regression models of the PDR did not reveal a statistically significant association with any co-variate, except male sex (Supplementary Figure 2), although the point estimate for the odds ratio for the I carrier x EPA interaction was consistent with colorectal polyp number findings (Figure 2).

## Discussion

A functional *FADS2* indel polymorphism (rs66698963) stratifies participants in the seAFOod trial that display colorectal polyp prevention by the *n*-3 HUFA EPA. This gene x treatment interaction was specific for EPA, as opposed to aspirin, compatible with current understanding of the mechanism of action of both agents. The elevated AA/LA ratio and higher AA-derived oxylipin metabolite levels in I allele carriers, support the biological plausibility of the association between one or more I alleles and increased colorectal polyp risk in untreated individuals, on one hand, and with polyp prevention efficacy of EPA, on the other.

The polyp prevention effect size for ‘sensitive’ I allele carriers is similar to that of aspirin, for which there is already a robust body of evidence supporting its anti-CRC activity [14]. It will now be important to validate rs66698963 as a CRC chemoprevention biomarker for EPA in independent cohorts, acknowledging that this was a *post hoc* trial analysis and the seAFOod polyp prevention trial cohort was predominantly White European and male [3], as well as determine whether rs66698963 genotype is predictive for other health benefits of EPA such as cardiovascular risk reduction [15].

## Data Availability

All data produced in the present study are available upon reasonable request to the authors

## Acknowledgements

The Authors wish to thank all the participants and research staff who contributed to the seAFOod polyp prevention trial. STOP-ADENOMA collaborator Dr Amy Downing (University of Leeds) provided helpful advice.

## Author contributions

The work reported in the paper has been performed by the authors, unless clearly specified in the text. KSDK, JTB, RB and MAH conceived and designed the study. MAH gained approvals for the study. GS, YNL, JD, RB, KSDK, JTB and MAH were involved in the acquisition, analysis, or interpretation of data. Drafting of the manuscript was done by GS, KSDK, JTB and MAH. MAH obtained funding for the study. All the authors contributed to the critical review and final approval of the manuscript. All authors were responsible for the decision to submit the manuscript. MAH had full access to all the data in the study and takes responsibility for the integrity of the data and the accuracy of the data analysis.

## Funding statement

The STOP-ADENOMA project (NIHR128210) was funded by the Efficacy and Mechanism Evaluation (EME) Programme, an MRC and NIHR partnership. The views expressed in this publication are those of the authors and not necessarily those of the MRC, NIHR or the Department of Health and Social Care. MAH is a NIHR Senior Investigator. MAH is supported by Cancer Research UK grant C23434/A24939.

## Data sharing statement

De-identified seAFOod trial meta-data are available upon request to the Chief Investigator and the trial Sponsor (University of Leeds).

## Conflict of interest disclosure

None of the Authors declare any potential conflict of interest.

## Notes

### Competing Interest Statement

The authors have declared no competing interest.

### Clinical Trial

ISRCTN05926847

### Funding Statement

This study was funded by the Efficacy and Mechanism Evaluation (EME) Programme (NIHR128210), an MRC and NIHR partnership. The views expressed in this publication are those of the authors and not necessarily those of the MRC, NIHR or the Department of Health and Social Care. MAH is a NIHR Senior Investigator. MAH is supported by Cancer Research UK grant C23434/A24939.

### Author Declarations

London and Surrey Borders Research Ethics Committee gave ethical approval for this work (19/LO/1655)

