## Supplementary material for "*FADS2* Indel polymorphism rs66698963 predicts colorectal polyp prevention by the *n*-3 fatty acid EPA"

**Ge Sun *et al*.**

### Supplementary Material


| | Page number |
|---|---|
| Methods | 2 |
| Figure 1 | 5 |
| Table 1 | 6 |
| Figure 2 | 7 |

**Supplementary Methods**

Laboratory methods

DNA extraction from buffy coat samples from seAFOod trial participants has been described previously [1]. Genotyping for the *FADS2* indel rs66698963 (I or D alleles) was performed by PCR on anonymized DNA samples not linked to trial data, as described [2, 3]. Baseline and on-treatment (6 months) RBC membrane HUFA levels (as the % of total fatty acids) and plasma oxylipin levels were measured by liquid chromatography-tandem mass spectrometry, as described and have been published [4, 5].

We calculated the product (AA)-precursor (LA) ratio in RBC membranes as a readout of FADS1 activity. The relationship between the AA/LA ratio and plasma total (*R*- and *S*- enantiomers) 15-HETE concentration with Indel genotype was tested using either the Wilcoxon or t-test, depending on normality testing.

Colorectal polyp outcomes in the seAFOod trial

The primary outcome of the seAFOod polyp prevention trial was the 'adenoma' (now more accurately termed 'polyp') detection rate (the % of individuals with one or more polyps (including adenomatous and serrated polyps) [4, 6]. We have argued that the secondary endpoint of polyp number is a more valid readout of polyp prevention efficacy in high-risk populations with multiple polyps [4, 6], and have since used polyp multiplicity, in parallel with the polyp detection rate (PDR) in secondary post-trial outcomes analyses [1, 5, 6].

Statistical analysis

The pre-specified analysis of colorectal polyp number in the original seAFOod trial report was performed by Poisson regression incorporating repeat colonoscopy at baseline in the trial and research site, at which the colonoscopy was undertaken, as co-variates [4]. We repeated this analysis for the smaller trial population (n=528) with an available Indel genotype, in order to confirm that the effect sizes for both EPA and aspirin were similar to the original randomised trial population [4]. However, the distribution of individual post-treatment total colorectal polyp counts better fits a negative binomial model [7], which has since been used in subsequent analyses of on-trial and post-trial colorectal polyp outcomes [1, 6]. Therefore, negative binomial regression was used to explore the interaction between treatment and rs66698963 genotype.

All analyses were conducted using R Studio, version 2021.09.0.

**Supplementary Methods references**

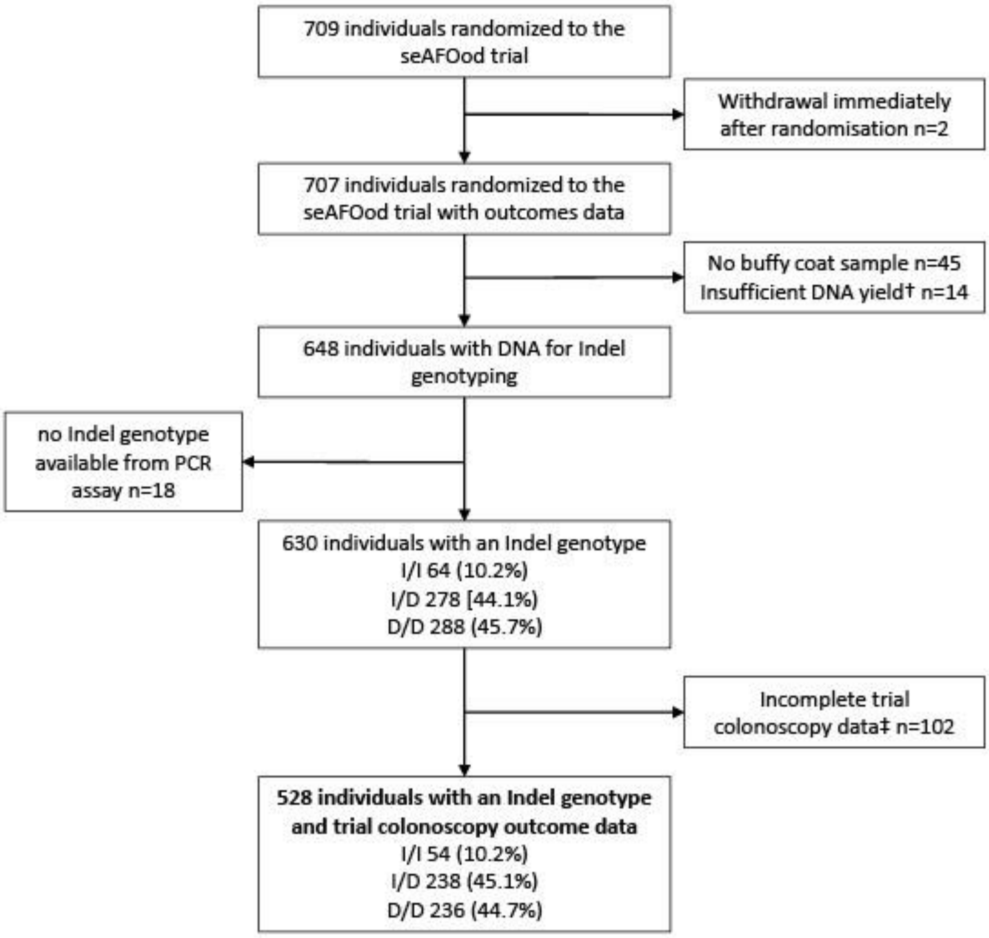


**Supplementary Figure 1. seAFOod trial participants and samples contributing to the treatment x Indel genotype analysis.** †DNA < 20 ng/ml. ‡Incomplete trial colonoscopy data includes 57 with no trial exit colonoscopy, 45 with missing data for repeat colonoscopy at baseline.

**Supplementary Table 1. Characteristics of 528 seAFOod trial participants with *FADS2* Indel genotype and trial colonoscopy outcome data**

| | Total study population | Placebos only | EPA only | Aspirin only | Aspirin & EPA | P for EPA *vs* no EPA | P for aspirin *vs* no aspirin | Excluded participants (incomplete colonoscopy data or no genotype) | P for included *vs* excluded groups |
|---|---|---|---|---|---|---|---|---|---|
| **Number of participants** | **528** | **138** | **123** | **136** | **131** | | | **179** | |
| **Age (median [IQR])** | 64.9 (6.3) | 64.5 (7.8) | 65.5 (6.2) | 64.8 (6.3) | 66.4 (7.7) | 0.3 | 0.2 | 66.4 (6.2) | 0.3 |
| **Sex** | | | | | | 0.5 | 0.6 | | 0.7 |
| Male | 423 (80.1)* | 109 (79.0) | 97 (78.9) | 107 (78.7) | 110 (84.0) | | | 140 (78.2) | |
| Female | 105 (19.9) | 29 (21.0) | 26 (21.1) | 29 (21.3) | 21 (16.0) | | | 39 (21.8) | |
| **Body mass index (BMI)†** | | | | | | 0.3 | 1.0 | | 0.9 |
| Underweight (<18.5) | 2 (0.4) | 0 | 1 (0.8) | 1 (0.7) | 0 | | | 1 (0.6) | |
| Normal (≥18.5 to <25 ) | 94 (17.9) | 23 (16.8) | 22 (18.2) | 18 (13.2) | 31 (23.7) | | | 30 (16.8) | |
| Overweight (≥25 to <30) | 228 (43.4) | 62 (45.3) | 49 (40.5) | 59 (43.4) | 58 (44.3) | | | 82 (45.7) | |
| Obese (≥30) | 201 (38.3) | 52 (37.9) | 49 (40.5) | 58 (42.6) | 42 (32.0) | | | 66 (36.9) | |
| **Diabetes** | | | | | | 0.4 | 0.1 | | 0.7 |
| No | 471 (89.2) | 118 (85.5) | 109 (88.6) | 123 (90.4) | 121 (92.4) | | | 155 (86.6) | |
| Yes | 57 (10.8) | 20 (14.5) | 14 (11.4) | 13 (9.6) | 10 (7.6) | | | 24 (13.4) | |
| **Tobacco smoking** | | | | | | 0.4 | 0.4 | | 0.5 |
| Never | 198 (37.5) | 49 (35.5) | 49 (39.8) | 50 (36.8) | 50 (38.2) | | | 56 (31.3) | |
| Ever | 252 (47.7) | 64 (46.4) | 66 (53.7) | 65 (47.8) | 57 (43.5) | | | 95 (53.1) | |
| Current | 78 (14.8) | 25 (18.1) | 8 (6.5) | 21 (15.4) | 24 (18.3) | | | 14 (15.6) | |
| **Alcohol intake†‡** | | | | | | 0.8 | 0.5 | | 0.8 |
| None | 80 (15.2) | 23 (16.7) | 19 (15.6) | 19 (14.0) | 19 (14.5) | | | 30 (16.9) | |
| 1-7 units/week | 179 (34.0) | 39 (28.3) | 47 (38.5) | 51 (37.5) | 42 (32.1) | | | 55 (31.2) | |
| 8-21 units/week | 161 (30.6) | 45 (32.6) | 29 (23.8) | 44 (32.4) | 43 (32.8) | | | 53 (29.9) | |
| ≥22 units/week | 107 (20.2) | 31 (22.5) | 27 (22.1) | 22 (16.2) | 27 (20.6) | | | 39 (22.0) | |

*figures in brackets are the % value unless indicated otherwise

†missing BMI value for one case in the placebo group and two cases in the EPA only group; missing alcohol intake data for one case in the EPA only group.

‡missing alcohol intake data for two cases in the excluded participant group.

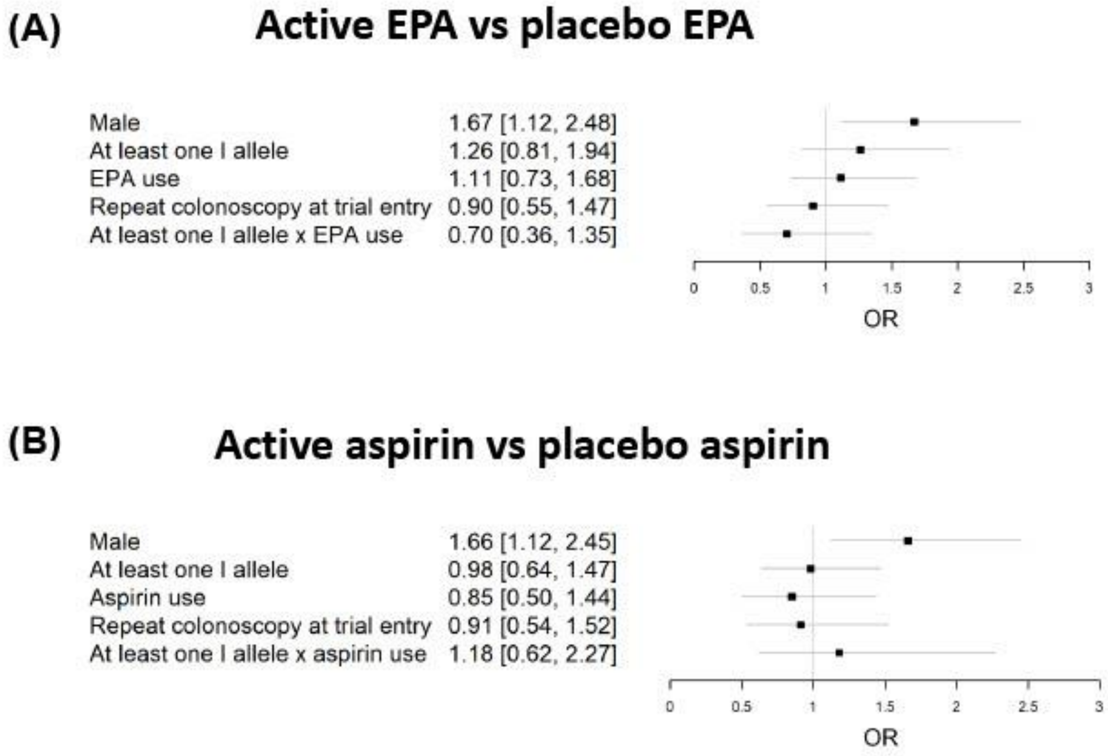


**Supplementary Figure 2. Multivariate models investigating the interaction between EPA and aspirin treatment, and FADS2 Indel genotype (I carrier status) for the polyp detection rate.** Forest plots with the odds ratio (OR) and 95% confidence interval for co-variates for 'at the margins' comparison of (A) active EPA *vs* placebo EPA, (B) active aspirin *vs* placebo aspirin.
